# Perceptions of Telerehabilitation Among Patients Receiving Physical Therapy for Spine Pain

**DOI:** 10.1101/2025.02.24.25322777

**Authors:** Kevin H. McLaughlin, Olivia Caan, Richard L. Skolasky

## Abstract

**Importance:** Telerehabilitation was widely used during the COVID-19 pandemic to maintain access to physical therapy. However, its use has declined since the pandemic, potentially due to changes in patient demand.

**Objective:** To explore patients’ beliefs, attitudes, and preferences regarding telerehabilitation for spine pain treatment.

**Design:** Email or telephone administered survey.

**Setting:** Outpatient physical therapy network affiliated with an academic healthcare system.

**Participants:** Patients recently evaluated for spine pain by a physical therapist (n=100).

**Main Outcomes and Measures:** Survey items addressed patients’ perceptions of telerehabilitation effectiveness, preferences for care delivery, barriers to telerehabilitation, and factors influencing telerehabilitation use.

**Results:** Patients viewed telerehabilitation as effective for treating spine pain but not as effective as in-clinic physical therapy. Patients reported a preference for receiving care in-clinic care or using a hybrid model, rather than receiving care through telerehabilitation alone. Barriers to telerehabilitation included a lack of awareness of who offers telerehabilitation and uncertainty about its effectiveness. Patients indicated that evidence supporting telerehabilitation effectiveness would increase their likelihood of using telerehabilitation in the future. Patients also reported that transportation challenges or increased COVID-19 rates would encourage them to use telerehabilitation in the future.

**Conclusions:** The results of this survey suggest that patients with spine pain believe telerehabilitation may be an effective treatment approach, but they are skeptical about using telerehabilitation instead of in-clinic physical therapy. These results also suggest that patients are open to hybrid approaches that incorporate telerehabilitation, as well as using telerehabilitation if circumstances prevented them from receiving their care in-clinic. The largest patient concern surrounding telerehabilitation seems to be the lack of evidence supporting its effectiveness.

**Relevance:** This study suggests that patients believe telerehabilitation will continue to play an important role post-pandemic. However, comparative effectiveness research is needed to address patient concerns surrounding the effectiveness of telerehabilitation and guide informed decision-making.

## Introduction

Changes in state-level practice acts for physical therapists, as well as changes in reimbursement, have facilitated the expansion of physical therapy delivered by telehealth, also referred to as telerehabilitation.^1^ Telerehabilitation stands to improve access to physical therapy by addressing barriers surrounding access (i.e., wait times, cost) and logistics (i.e., missed work time, transportation).^2-7^ Early evidence also suggests that telerehabilitation may be an effective treatment approach for patients with health conditions commonly treated by physical therapists, including spine pain.^8-10^

Despite its advantages, the use of telerehabilitation has declined substantially since the early stages of the pandemic.^1^ There are a number of factors that likely influence these utilization rates, including patient and provider perceptions of telerehabilitation. A survey of orthopaedic physical therapists conducted after the end of the COVID-19 Public Health Emergency suggests that physical therapists consider telerehabilitation to be an effective treatment approach for patients with musculoskeletal pain but do not consider telerehabilitation to be as effective as in-clinic physical therapy.^11^ If physical therapists consider telerehabilitation to be inferior to in-clinic care, they may be less likely to offer these services, which would decrease the availability (i.e., supply) of telerehabilitation services. Likewise, patients’ perceptions of telerehabilitation are likely to influence the demand for these services. However, no studies have examined patients’ perceptions of telerehabilitation since the end of the COVID-19 pandemic.

To address this gap in knowledge, we conducted a survey of patients seeking physical therapy care for spine pain. The purpose of this study was to examine patients’ perceptions of telerehabilitation, including perceptions of clinical effectiveness, barriers to utilizing telerehabilitation, and factors that would influence their use of telerehabilitation in the future.

## Methods

### Survey Design

An online survey was developed for the purposes of this study by members of the study team with expertise in physical therapy and telerehabilitation. The survey included 23 questions separated into 5 sections: (1) Previous experience using physical therapy and telehealth, (2) perceptions and preferences for telerehabilitation, (3) barriers to using telerehabilitation, (4) factors affecting future use of telerehabilitation, and (5) respondent characteristics. Survey items included a combination of multiple choice and Likert-style responses. The full version of the survey is available in Supplemental Material.

### Survey Distribution

We used a convenience sample of adult patients (>18 years) evaluated for spine pain by an outpatient physical therapist at our institution in the previous 6 months. Patients meeting these criteria were initially emailed an invitation to participate in the study, which included a link to the online survey. Patients not responding to our email invitation within 7 days were called by research coordinators, who conducted the survey over the phone or provided a new email invitation with a link to the online survey. The survey was administered using REDCap.^12^

### Data Analysis

Summary statistics were used to describe survey responses, including the number and proportion of patients that selected each survey response option.

### Ethics Approval

This study was recognized as exempt by the Johns Hopkins Medicine Institutional Review Board (IRB00420821).

## Results

### Respondent Characteristics

We received a total of 100 complete survey responses. Age of respondents ranged from 29 to 87 years, with those over 65 representing the most common age category (n=38, 38%). Respondents were majority non-Hispanic (n=94, 94%) and identified most commonly as White (n=65, 65%) or Black (n=27, 27%). Most patients lived in suburban areas (n=60, 60%), followed by urban areas (n=28, 28%) and rural areas (n=7, 7%). Over half of respondents had previously used some form of telehealth (n=56, 56%), but only 8% (n=8) reported using telerehabilitation in the past.

### Perceptions and Preferences for Telerehabilitation

Figure 1 describes patients’ responses to statements surrounding the effectiveness of telerehabilitation. Patients’ responses to a statement indicating that telerehabilitation could be an effective approach for patients’ current condition were mixed. Thirty percent (n=30) and 13% (n=13) of patients indicated that they “agree” or “strongly agree” with this statement, while 25% (n=25) and 15% (n=15) of patients disagreed and strongly disagreed with this statement, respectively. Seventeen percent (n=17) of patients reported a neutral response. In response to a statement indicating that telerehabilitation would be as effective as in-clinic care for their current condition, roughly two-thirds of patients indicated that they “disagree” (n=41, 41%) or strongly disagreed (n=23, 23%). In response to a statement suggesting that telerehabilitation would be more effective than in-clinic care for their current condition, a similar proportion of patients indicated they “disagree” (n=40, 40%) or “strongly disagree” (n=35, 35%).

**Figure 1.**
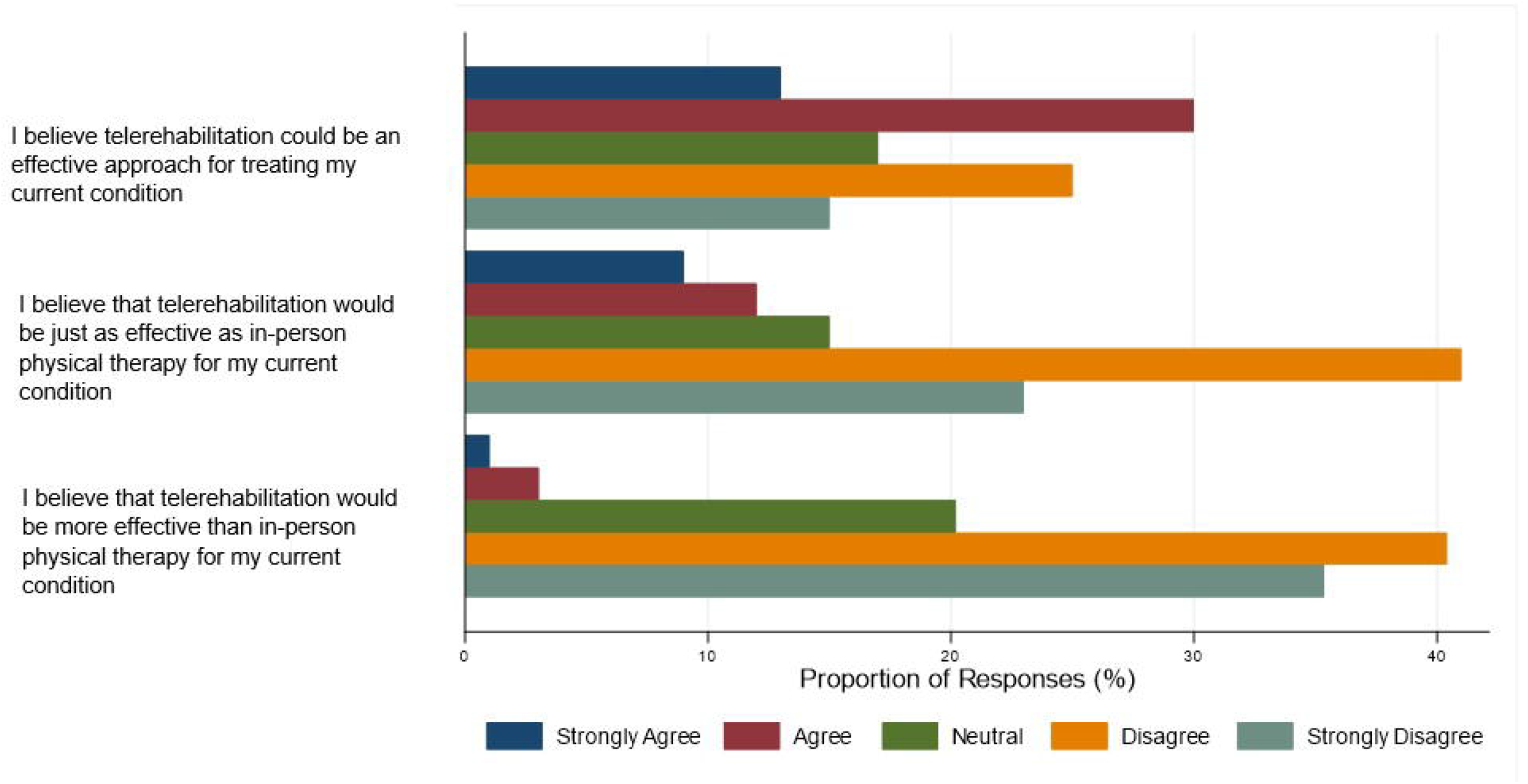

Figure 2 describes patients’ responses to statements about the way(s) they prefer to receive physical therapy care. In response to a statement saying the patient would prefer to receive all physical therapy care in-clinic, most participants indicated that they strongly agree (n=42, 42%) or agree (n=27, 27%). In response to a statement saying that the patient would prefer to receive physical therapy care using a combination of in-clinic visits and telerehabilitation, the most common response was “agree” (n=36, 36%). In response to a statement saying that the patient would prefer to receive all visits using telerehabilitation, roughly 9 out of 10 patients indicated they “disagree” (n=51, 51%) or “strongly disagree” (n=36, 36%). In response to a statement indicating that patients would be open to using telerehabilitation if circumstances prevented them from attending physical therapy in-clinic, most patients responded saying they “agree” (n=46, 46%) or “strongly agree” (n=28, 28%).

**Figure 2.**
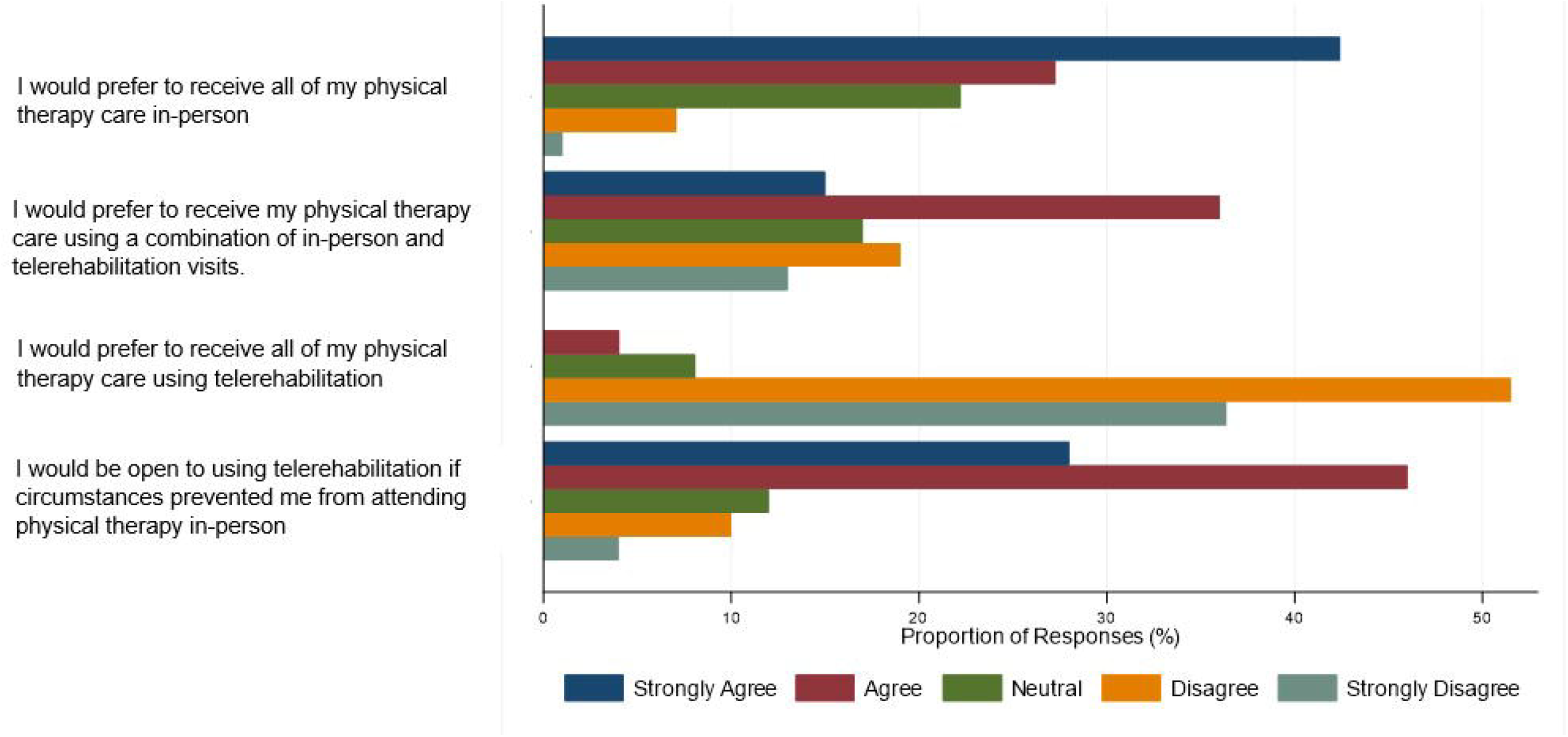

### Barriers to Using Telerehabilitation

Figure 3 describes patients’ responses to statements about barriers to using telerehabilitation. Most patients indicated that internet access (n=89, 89%); access to a computer, tablet, or smartphone (n=89, 89%); comfortability using technology (n=78, 78%); or a lack of privacy at home (n=86, 86%) were “not a barrier” to using telerehabilitation. Over 50% of patients reported that not knowing who offers telerehabilitation and or not knowing if telerehabilitation is effective for their condition were at least somewhat of a barrier to them using telerehabilitation. Slightly over half (n=52, 53%) of patients reported that they did not consider insurance coverage a barrier to using telerehabilitation, with the remaining patients (48%) reporting that insurance coverage was at least somewhat of a barrier to using telerehabilitation.

**Figure 3.**
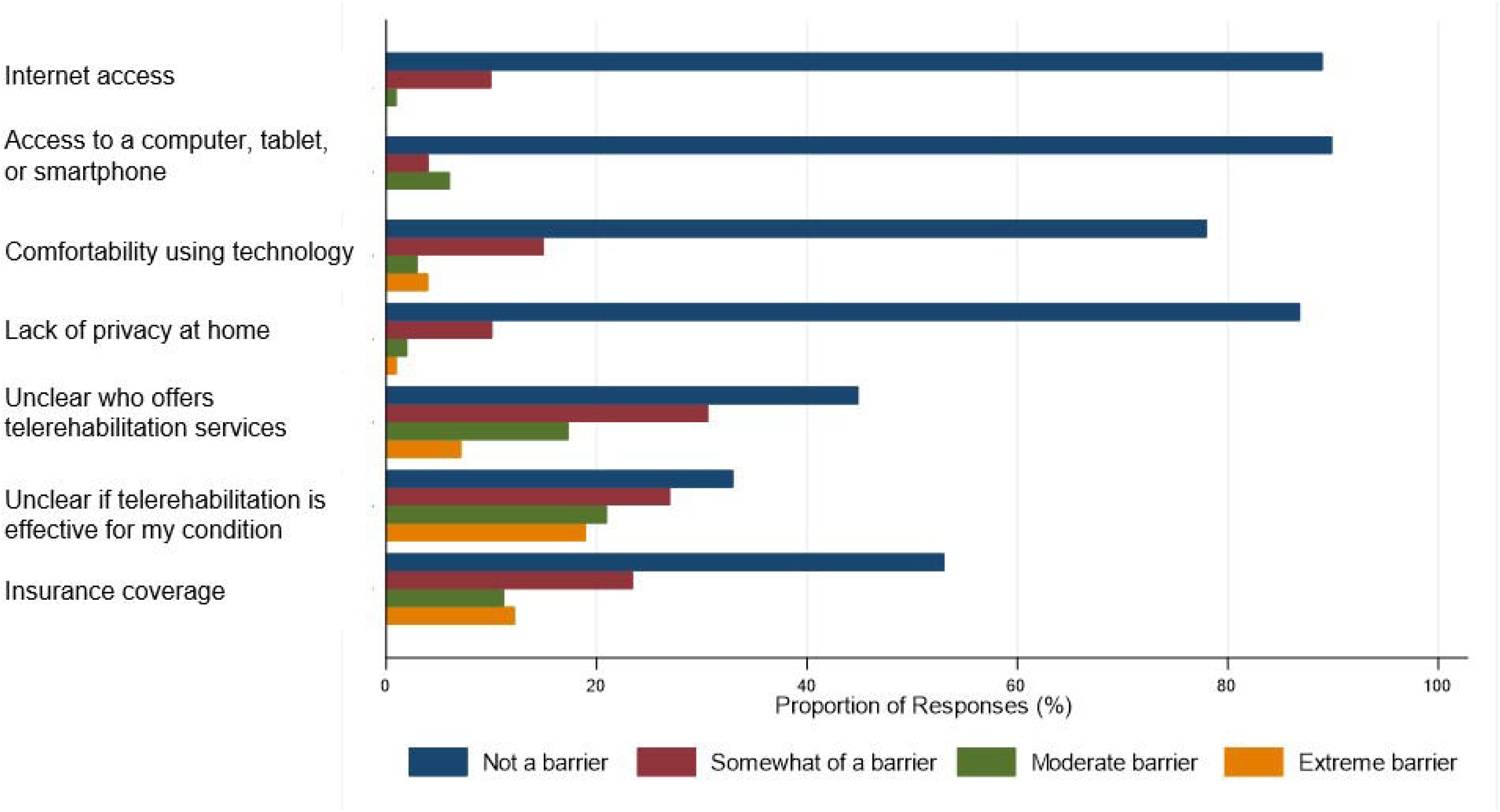

### Factors Affecting Future Telerehabilitation Use

Figure 4 describes patients’ responses to statements about how specific factors might influence their use of telerehabilitation in the future. Patients indicated that difficulty accessing transportation (n=45, 45%), a rise in COVID infection rates (n=50, 50%), or new research supporting the effectiveness of telerehabilitation (n=57, 57%) would make them more likely to use telerehabilitation. Half of patients (n=50, 50%) reported that decreased wait times would not influence their use of telerehabilitation, with most other patients (n=45, 45%) indicating that decreased wait times would make them more likely to use telerehabilitation. Discounted co-payments were reported to have no influence on the likelihood of using telerehabilitation by most patients (n=65, 65%).

**Figure 4.**
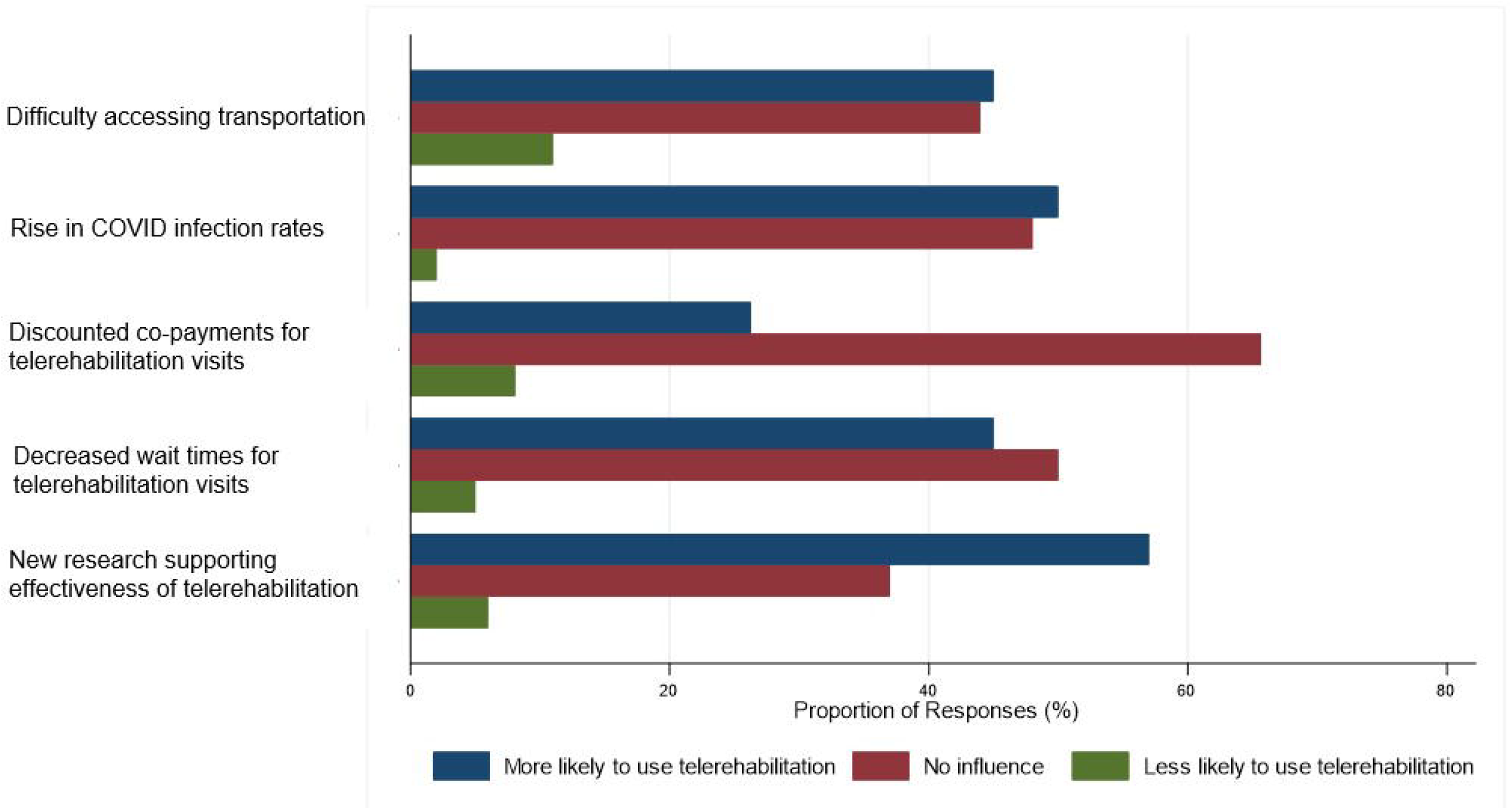

## Discussion

This study utilized a survey to examine attitudes and beliefs surrounding telerehabilitation among patients who have recently seen a physical therapist for spine pain. A large proportion of patients surveyed in this study indicated that they believe telerehabilitation could be an effective treatment approach for their condition. However, the majority of surveyed patients reported that they did not believe telerehabilitation would be as effective as in-clinic care for their condition and preferred to receive their care entirely in-clinic or using a hybrid approach that includes in-clinic and telerehabilitation visits. Patients largely agreed that they would be open to using telerehabilitation if circumstances prevented them from attending in-clinic physical therapy and they would be more likely to use telerehabilitation in the future if new research findings supported its effectiveness.

The results of this study align with prior research in this area. In a previous interventional cohort study (n=31), patients receiving telerehabilitation for chronic low back pain during the COVID-19 pandemic reported benefits of telerehabilitation that included convenience, time savings, and personalized 1-on-1 care.^13^ At the same time, patients had concerns about the quality of care that physical therapists can deliver through telerehabilitation and the lack of hands-on care inherent to telerehabilitation. Like patients surveyed in the current study, those in the previous study recommended a hybrid approach to physical therapy that includes in-clinic evaluation and a combination of in-clinic and virtual follow ups. Another study used a national survey of patients with orthopaedic conditions (n=61) to explore patients’ attitudes and beliefs surrounding telerehabilitation during the pandemic.^14^ Similar to our study, patients surveyed as part of the previous study reported telerehabilitation to be accessible, unencumbered by access or familiarity with the internet and technology. The most common concern among patients in the previous study was if the quality of care that can be delivered using telerehabilitation is inferior to in-clinic care. The results of the current study build upon the results of previous studies by re-examining patients’ perceptions of telerehabilitation following the pandemic and restrictions on in-clinic physical therapy visits. Our results also provide more granular insights into the factors that might influence future decisions to use telerehabilitation.

The results of this study suggest that patients’ perceptions of telerehabilitation are similar to physical therapists’ perceptions of telerehabilitation. Our study team recently conducted a survey of orthopaedic physical therapists about their perceptions of telerehabilitation for patients with musculoskeletal conditions.^11^ The results of the provider survey indicated that physical therapists consider telerehabilitation to be an effective method of care delivery for patients with musculoskeletal pain, but do not consider telerehabilitation to be as effective as in-clinic care for this population. They also reported that they would prefer to use telerehabilitation as part of a hybrid program (i.e., combination of in-clinic and telerehabilitation visits) instead of providing all of a patient’s visits through telerehabilitation. The results of this patient survey suggest that patients and providers agree in these areas.

The results of this study may inform telerehabilitation strategies for healthcare systems and individual physical therapy clinics. For example, this study suggests that only a small proportion of patients attending physical therapy would prefer to receive all of their care using telerehabilitation. However, a much higher proportion of patients reported that they would be interested in receiving their care through a combination of in-clinic and telerehabilitation visits. Taken together, these results suggest that patients may be interested in using telerehabilitation as part of a hybrid care model versus a telerehabilitation-only model. Patients also suggested that they would be open to receiving care using telerehabilitation if circumstances prevented them from attending in-clinic. Providing patients with the chance to convert in-clinic visits to telerehabilitation visits may provide an opportunity to reduce cancellations and no-shows in the event that patients become sick, have last-minute scheduling conflicts, etc.

Over half of survey respondents (57%) in this study reported that they would be more willing to use telerehabilitation if research emerged that supported its effectiveness for their condition. At the time of this publication, research on the effectiveness of telerehabilitation for spine pain is limited. Most studies in this area have primarily focused on self-management approaches, such as mobile applications, versus care provided using real-time video visits with a physical therapist.^15,16^ Results from studies examining the use of physical therapy delivered using real-time video visits suggest that this approach may improve pain and disability among patients with spine pain.^9,10^ However, these studies have utilized observational or single-arm study designs, which do not allow for conclusions to be drawn regarding the effectiveness of telerehabilitation compared to in-clinic physical therapy. Studies examining the comparative effectiveness of in-clinic physical therapy and telerehabilitation are needed to inform patient treatment decisions regarding how they prefer to receive physical therapy for spine pain. Moreover, results from these studies need to be disseminated through patient-facing resources (e.g., social media), in addition to more traditional provider-facing resources (e.g., peer-review journals, academic conferences).

It is important to consider the characteristics of the patient sample included in this study when interpreting our survey results. This study included a convenience sample of individuals who had recently completed a physical therapy evaluation for spine pain. All of the patients included in this study attended at least 1 in-clinic physical therapy visit. As such, the results of this study are likely most reflective of the attitudes and beliefs surrounding telerehabilitation among patients with the ability to receive care either in-clinic or by telerehabilitation. This represents a large population reflective of the populations served by many healthcare systems and individual physical therapy clinics. However, our results may not reflect the perceptions of telerehabilitation held by patients without access to in-clinic physical therapy, such as those in rural or remote parts of the country. In an observational cohort of patients with chronic low back pain receiving telerehabilitation during the COVID-19 pandemic, a qualitative study revealed that most patients had a positive or neutral experience with telerehabilitation and, compared to those with a negative experience, endorsed higher therapeutic bond with their physical therapist.^13^ It is possible that patients with less access to in-clinic physical therapy have different perceptions and preferences surrounding telerehabilitation.

Limitations of this study include its single-institution design. It is possible that the perceptions of telerehabilitation held by patients receiving care at our institution may not be reflective of patients seen at other institutions. Our study results have also been affected by response bias, as patients with stronger opinions on telerehabilitation may have been more likely to respond to our study invitation.

## Conclusion

Patients consider telerehabilitation to be a potentially effective treatment strategy for painful spinal conditions, but do not believe that telerehabilitation is as effective as in-clinic physical therapy. Patients reported they are open to hybrid programs that include in-clinic and telerehabilitation visits, as well as using telerehabilitation in the event that circumstances prevent them from attending physical therapy in person. Patients also reported that they would be more likely to use telerehabilitation if evidence emerged supporting its effectiveness.

## Data Availability

All data produced in the present study are available upon reasonable request to the authors

---

**Table 1.** Participant Characteristics.

|  |  |
| --- | --- |
| **Age, N (%)** |  |
| 25-49 | 20 (20) |
| 50-64 | 37 (39) |
| 65+ | 38 (38) |
| Not answered | 5 (5) |
| **Female** | 62 (62) |
| **Race, N (%)** |  |
| American Indian or Native Alaskan | 1 (1) |
| Asian | 6 (6) |
| Black or African American | 27 (27) |
| White or Caucasian | 65 (65) |
| Not answered | 1 (1) |
| **Ethnicity, N (%)** |  |
| Hispanic | 4 (4) |
| Non-Hispanic | 94 (94) |
| Not answered | 2 (2) |
| **Area of residence, N (%)** |  |
| Urban | 28 (28) |
| Suburban | 60 (60) |
| Rural | 7 (7) |
| **Previously used any kind of telehealth, N (%)** | 56 (56) |
| **Previously used telehealth physical therapy, N (%)** | 8 (8) |

## Literature Cited

1. McLaughlin KH, Levy JF, Fritz JM, Skolasky RL. Trends in Telerehabilitation Utilization in the United States 2020-2021. Archives of Physical Medicine and Rehabilitation. 2024;

2. Carter SK, Rizzo JA. Use of outpatient physical therapy services by people with musculoskeletal conditions. Physical therapy. 2007;87(5):497–512.

3. Carvalho E, Bettger JP, Goode AP. Insurance Coverage, Costs, and Barriers to Care for Outpatient Musculoskeletal Therapy and Rehabilitation Services. North Carolina Medical Journal. September 1, 2017 2017;78(5):312. doi:10.18043/ncm.78.5.312

4. Castillo RC, MacKenzie EJ, Webb LX, Bosse MJ, Avery J. Use and perceived need of physical therapy following severe lower-extremity trauma. Archives of physical medicine and rehabilitation. 2005 Sep 2005;86(9):1722–1728. doi:10.1016/j.apmr.2005.03.005

5. Dolot J, Viola D, Shi Q, Hyland M. Impact of Out-of-Pocket Expenditure on Physical Therapy Utilization for Nonspecific Low Back Pain: Secondary Analysis of the Medical Expenditure Panel Survey Data. Physical Therapy. February 1, 2016 2016;96(2):212–221. doi:10.2522/ptj.20150028

6. Dolot J, Hyland M, Shi Q, Kim H-Y, Viola D, Hoekstra C. Factors Impacting Physical Therapy Utilization for Patients With Nonspecific Low Back Pain: Retrospective Analysis of a Clinical Data Set. Physical Therapy. August 31, 2020 2020;100(9):1502–1515. doi:10.1093/ptj/pzaa082

7. Jette AM. The Promise and Potential of Telerehabilitation in Physical Therapy. Physical Therapy. 2021-03-03 2021;101(3):pzab081. doi:10.1093/ptj/pzab081

8. Cottrell MA, O’Leary SP, Raymer M, Hill AJ, Comans T, Russell TG. Does telerehabilitation result in inferior clinical outcomes compared with in-person care for the management of chronic musculoskeletal spinal conditions in the tertiary hospital setting? A non-randomised pilot clinical trial. Journal of Telemedicine and Telecare. 2021;27(7):444–452. doi:10.1177/1357633×19887265

9. Fritz JM, Minick KI, Brennan GP, et al. Outcomes of Telehealth Physical Therapy Provided Using Real-Time, Videoconferencing for Patients With Chronic Low Back Pain: A Longitudinal Observational Study. Archives of Physical Medicine and Rehabilitation. 2022/06/03/ 2022;doi:10.1016/j.apmr.2022.04.016

10. Werneke MW, Deutscher D, Hayes D, Grigsby D, Mioduski JE, Resnik LJ. Is Telerehabilitation a Viable Option for People With Low Back Pain? Associations Between Telerehabilitation and Outcomes During the COVID-19 Pandemic. Phys Ther. May 5 2022;102(5)doi:10.1093/ptj/pzac020

11. McLaughlin K, Minick KI, Fritz JM, et al. Physical therapists’ perceptions of telerehabilitation for patients with musculoskeletal conditions in a post-pandemic world. medRxiv (pre-print). 2025:2025.01.17.25320739. doi:10.1101/2025.01.17.25320739

12. Harris PA, Taylor R, Minor BL, et al. The REDCap consortium: Building an international community of software platform partners. Journal of Biomedical Informatics. Jul 2019 2019;95:103208. doi:10.1016/j.jbi.2019.103208

13. Skolasky RL, Kimball ER, Galyean P, et al. Identifying Perceptions, Experiences, and Recommendations of Telehealth Physical Therapy for Patients with Chronic Low Back Pain: A Mixed Methods Survey. Archives of Physical Medicine and Rehabilitation. 2022/07/06/ 2022;doi:https://doi.org/10.1016/j.apmr.2022.06.006

14. Saaei F, Klappa SG. Rethinking Telerehabilitation: Attitudes of Physical Therapists and Patients. J Patient Exp. 2021;8:23743735211034335. doi:10.1177/23743735211034335

15. Tabacof L, Baker TS, Durbin JR, et al. Telehealth treatment for nonspecific low back pain: A review of the current state in mobile health. PM&R. 2022;doi:10.1002/pmrj.12738

16. Nicholl BI, Sandal LF, Stochkendahl MJ, et al. Digital Support Interventions for the Self-Management of Low Back Pain: A Systematic Review. Journal of Medical Internet Research. 2017-5-21 2017;19(5)doi:10.2196/jmir.7290

