## Supplemental Material for "Perceptions of Telerehabilitation Among Patients Receiving Physical Therapy for Spine Pain"

**Patient Perceptions of Telerehabilitation for Musculoskeletal Pain Post-Pandemic**

The use of telehealth expanded greatly in the US during the COVID-19 pandemic. However, it is unclear how telehealth services can and should be used after the end of the pandemic. This survey is being conducted to examine patients’ attitudes and beliefs surrounding care delivered by telehealth, also referred to as *telerehabilitation*. This survey takes approximately 6 minutes to complete.

Completion of this survey will serve as your consent to participate in this study. All responses will be collected anonymously.

Please note that in this survey, the term *telerehabilitation* is used to refer to physical therapy care delivered remotely, using real-time video visits.

***Section 1: Telerehabilitation Experience Before, During and After the COVID-19 Pandemic***

*For each question, please select the response that most closely describes your previous or current use of telerehabilitation.*

1. Excluding care that you are currently receiving, have you received physical therapy in the past?
   a. Yes
   b. No
2. IF YES: How did you receive physical therapy care?
   a. In-person
   b. Telerehabilitation (i.e., video or phone visits)
   c. Combination of the above
3. Have you ever received telehealth services from another type of health care provider?
   a. Yes
   b. No
4. IF YES: What type of provider? [free text]

***Section 2: Perceptions of Telerehabilitation***

*Please indicate your level of agreement with each of the following statements.*

5. I believe telerehabilitation could be an effective approach for treating my current condition
   a. Strongly agree (5)
   b. Agree (4)
   c. Neutral (3)
   d. Disagree (2)
   e. Strongly disagree (1)
6. I believe that telerehabilitation would be just as effective as in-person physical therapy for my current condition.
   a. Strongly agree (5)
   b. Agree (4)
   c. Neutral (3)
   d. Disagree (2)
   e. Strongly disagree (1)
7. I believe that telerehabilitation would be more effective than in-person physical therapy for my current condition.
   a. Strongly agree (5)
   b. Agree (4)
   c. Neutral (3)
   d. Disagree (2)
   e. Strongly disagree (1)
8. I would prefer to receive all of my physical therapy care in-person.
   a. Strongly agree (5)
   b. Agree (4)
   c. Neutral (3)
   d. Disagree (2)
   e. Strongly disagree (1)
9. I would prefer to receive my physical therapy care using a combination of in-person and telerehabilitation visits.
   a. Strongly agree (5)
   b. Agree (4)
   c. Neutral (3)
   d. Disagree (2)
   e. Strongly disagree (1)
10. I would prefer to receive all of my physical therapy care using telerehabilitation.
    a. Strongly agree (5)
    b. Agree (4)
    c. Neutral (3)
    d. Disagree (2)
    e. Strongly disagree (1)
11. I would be open to using telerehabilitation if circumstances prevented me from attending physical therapy in-person.
    a. Strongly agree (5)
    b. Agree (4)
    c. Neutral (3)
    d. Disagree (2)
    e. Strongly disagree (1)

***Section 3: Barriers to Telerehabilitation***

*Please indicate your level of agreement with each of the following statements.*

12. Please indicate to what extent each of the following items is a barrier to your use of telerehabilitation

|  | **Not a barrier** | **Somewhat of a barrier** | **Moderate barrier** | **Extreme barrier** |
| --- | --- | --- | --- | --- |
| Internet access |  |  |  |  |
| Access to a computer, tablet, or smart phone |  |  |  |  |
| Comfortability using technology |  |  |  |  |
| Lack of privacy at home |  |  |  |  |
| Unclear who offers telerehabilitation services |  |  |  |  |
| Unclear if telerehabilitation is effective for my condition |  |  |  |  |
| Insurance coverage |  |  |  |  |

13. Are there any other barriers that affect your ability to receive telerehabilitation? [Free text]

***Section 4. Factors Affecting Use of Telerehabilitation***

14. Please indicate how each of the following scenarios or details would influence the likelihood of you attending telerehabilitation instead of in-person physical therapy in the future

|  | **More likely to use telerehabilitation** | **Neutral** | **Less likely to use telerehabilitation** |
| --- | --- | --- | --- |
| Difficulty accessing transportation |  |  |  |
| A rise in COVID infection rates |  |  |  |
| Discounted co-payments for telerehabilitation visits |  |  |  |
| Decreased wait time for telerehabilitation visits |  |  |  |
| New research shows telerehabilitation is effective for your condition |  |  |  |

15. Please enter anything that would increase or decrease your willingness to receive telerehabilitation not included above: [free text]

***Section 5: Respondent Characteristics***

16. Which of the following best describes the region you live in?
    a. a. South Atlantic (DE, MD, DC, GA, NC, PR, SC, VA, WV, FL)
    b. Middle Atlantic (NJ, NY, PA)
    c. East North Central (IL, MI, OH, WI)
    d. West North Central (IA, KS, MN, MO, NE, ND, SD)
    e. East South Central (AL, KY, MS, TN)
    f. West South Central (AR, LA, OK, TX)
    g. New England (CT, ME, MA, NH, RI, VT)
    h. Pacific (AK, CA, HI, OR, WA)
    i. Mountain (AZ, CO, ID, MT, NV, NM, UT, WY)
17. Which of the following best describes the area that you live in?
    a. Urban
    b. Suburban
    c. Rural

18. How do you describe yourself?
    a. Female
    b. Male
    c. Non-binary/third gender
    d. Prefer to self-describe: ______________________
    e. Prefer not to say
19. What race best describes you?
    a. American Indian or Native Alaskan
    b. Asian
    c. Black or African American
    d. Native Hawaiian or Pacific Islander
    e. White or Caucasian
20. What ethnicity best describes you?
    a. Hispanic
    b. Non-Hispanic
21. How old are you? ____ years
22. What is the highest level of education you have completed?
    a. Less than high school
    b. High school or GED
    c. Some College
    d. Advanced degree (Masters, Doctorate)
23. What type of insurance do you have?
    a. Medicare
    b. Medicaid
    c. Commercial insurance
    d. Not insured
